# Structural neuroimaging biomarkers for obsessive-compulsive disorder in the ENIGMA-OCD consortium: medication matters

**DOI:** 10.1101/19012567

**Authors:** Willem B. Bruin, Luke Taylor, Rajat M. Thomas, Jonathan P Shock, Paul Zhutovsky, Yoshinari Abe, Pino Alonso, Stephanie H. Ameis, Alan Anticevic, Paul D. Arnold, Francesca Assogna, Francesco Benedetti, Jan C. Beucke, Premika S.W. Boedhoe, Irene Bollettini, Anushree Bose, Silvia Brem, Brian P. Brennan, Jan K Buitelaar, Rosa Calvo, Yuqi Cheng, Kang Ik K. Cho, Sara Dallaspezia, Damiaan Denys, Benjamin A. Ely, Jamie D. Feusner, Kate D. Fitzgerald, Jean-Paul Fouche, Egill A. Fridgeirsson, Patricia Gruner, Deniz A. Gürsel, Tobias U. Hauser, Yoshiyuki Hirano, Marcelo Q. Hoexter, Hao Hu, Chaim Huyser, Iliyan Ivanov, Anthony James, Fern Jaspers-Fayer, Norbert Kathmann, Christian Kaufmann, Kathrin Koch, Masaru Kuno, Gerd Kvale, Jun Soo Kwon, Yanni Liu, Christine Lochner, Luisa Lázaro, Paulo Marques, Rachel Marsh, Ignacio Martínez-Zalacaín, David Mataix-Cols, José M. Menchón, Luciano Minuzzi, Pedro S. Moreira, Astrid Morer, Pedro Morgado, Akiko Nakagawa, Takashi Nakamae, Tomohiro Nakao, Janardhanan C. Narayanaswamy, Erika L. Nurmi, Joseph O’Neill, Jose C. Pariente, Chris Perriello, John Piacentini, Fabrizio Piras, Federica Piras, Y.C. Janardhan Reddy, Oana G. Rus-Oswald, Yuki Sakai, João R. Sato, Lianne Schmaal, Eiji Shimizu, H. Blair Simpson, Noam Soreni, Carles Soriano-Mas, Gianfranco Spalletta, Emily R. Stern, Michael C. Stevens, S. Evelyn Stewart, Philip R. Szeszko, David F. Tolin, Ganesan Venkatasubramanian, Zhen Wang, Je-Yeon Yun, Daan van Rooij, ENIGMA-OCD consortium, Paul M. Thompson, Odile A. van den Heuvel, Dan J. Stein, Guido A. van Wingen

**Author notes:** **corresponding author.** Postal address: Meibergdreef 5,1105 AZ, Amsterdam, The Netherlands. see excel sheet for affiliations and full consortium member authors.

## Abstract

**Objective:** No diagnostic biomarkers are available for obsessive-compulsive disorder (OCD). Magnetic resonance imaging (MRI) studies have provided evidence for structural abnormalities in distinct brain regions, but effect sizes are small and have limited clinical relevance. To investigate whether individual patients can be distinguished from healthy controls, we performed multivariate analysis of structural neuroimaging data from the ENIGMA-OCD consortium.

**Method:** We included 46 data sets with neuroimaging and clinical data from adult (≥18 years) and pediatric (<18 years) samples. T_1_ images from 2,304 OCD patients and 2,068 healthy controls were analyzed using standardized processing to extract regional measures of cortical thickness, surface area and subcortical volume. Machine learning classification performance was tested using cross-validation, and possible effects of clinical variables were investigated by stratification.

**Results:** Classification performance for OCD versus controls using the complete sample with different classifiers and cross-validation strategies was poor (AUC—0.57 (standard deviation (SD)=0.02;P_corr_=0.19) to 0.62 (SD=0.03;P_corr_<.001)). When models were validated on completely new data from other sites, model performance did not exceed chance-level (AUC—0.51 (SD=0.11;P_corr_>.99) to 0.54 (SD=0.08;P_corr_>.99)). In contrast, good classification performance (>0.8 AUC) was achieved within subgroups of patients split according to their medication status.

**Conclusions:** Parcellated structural MRI data do not enable good distinction between patients with OCD and controls. However, classifying subgroups of patients based on medication status enables good identification at the individual subject level. This underlines the need for longitudinal studies on the short- and long-term effects of medication on brain structure.

## Introduction

Obsessive-compulsive disorder (OCD) is a severe and disabling condition that occurs in 2-3% of the population [1]. It is characterized by recurrent, intrusive, irrational and distressing thoughts (obsessions) and repetitive behaviors or mental acts (compulsions) [2]. So far, no biomarkers that may aid differential diagnosis are available, and diagnosis relies entirely on recognition of behavioral features assessed by clinical interview [3]. Many neuroimaging studies have provided evidence for abnormalities in cortico-striato-thalamo-cortical (CSTC) circuits, as well as distributed changes in limbic, parietal and cerebellar regions [4, 5]. This has recently been confirmed by meta- and mega-analysis of neuroimaging studies within the Enhancing Neuro-Imaging and Genetics through Meta-Analysis (ENIGMA) consortium [6–8]. However, inference was done at the group-level, and the small effect sizes that were reported precludes clinical application.

Analytic tools such as multivariate pattern analysis (MVPA) enable inference at the individual-level, which may result in better discrimination [3, 9]. MVPA techniques can be used to develop predictive models that extract common patterns from neuroimaging data to classify individuals based on their diagnosis. A major advantage of MVPA is its ability to use inter-regional correlations to detect subtle and spatially distributed effects compared to traditional methods of analysis [4]. Therefore, MVPA seems particularly suited for neuroimaging analyses in OCD, as abnormalities are typically distributed across the brain [10, 11]. Previous MVPA studies have been able to distinguish OCD patients from controls with accuracies ranging from 66-100% (reviewed in [12]). Although these results are promising, sample sizes have typically been small, limiting model performance optimization and leading to high variance in estimated accuracy and overly optimistic classification rates [13]. Additionally, most studies have been performed using data from one research center to minimize technical (e.g., scanner hardware, protocols and diagnostic assessment) and clinical (e.g., age, medication status, disease chronicity and severity) heterogeneity. It is therefore not clear whether these results generalize to other centers, which would be required for clinical application [14–16].

Here, we used data from the ENIGMA-OCD consortium, including 4,372 participants recruited at 36 research institutes around the world, with a full range of technical and clinical heterogeneity. We assessed the ability of MVPA to distinguish OCD patients from healthy controls based using structural neuroimaging data at the individual subject level. We investigated machine learning classification performance in both single-site and multi-site samples using different validation strategies to assess generalizability, as well as effects of clinical variables, such as medication use.

## Materials and methods

### Study population

The ENIGMA-OCD working group includes 46 data sets from 36 international research institutes, with neuroimaging and clinical data from adult (≥18 years) and pediatric (<18 years) samples. In total, we analyzed data from 4,372 participants, including 2,304 OCD patients (n=1,801 adult, n=503 child) and 2,068 healthy controls (HC; i.e., free of psychopathology; n=1,629 adult, n=439 child), with 38 of 46 datasets identical to those described in previous mega-analyses by this working group [6, 7, 17]. All participating sites obtained permission from their local institutional review boards or ethics committees to provide anonymized data for analysis, and all study participants provided written informed consent. Demographic and clinical characteristics of each cohort are detailed in supplementary **Table S1**. A complete overview of instruments used to obtain diagnosis and clinical information can be found elsewhere (Data Supplement 1, Supplementary Section S1) [7]. Diagnosis was determined in accordance with DSM [2]; MINI and SCID were used for adult samples and K-SADS, MINI-KIDS and ADIS were used for pediatric samples [18–22].

### MRI processing

Structural T1-weighted brain MRI scans were acquired and processed locally at each site. Image acquisition parameters are listed elsewhere[7]. Parcellations were performed using FreeSurfer (FS) software version 5.3 (http://surfer.nmr.mgh.harvard.edu), following standardized ENIGMA protocols to harmonize analyses and quality control procedures across multiple sites (see http://enigma.usc.edu/protocols/imaging-protocols/). Parcellations of 34 cortical (Desikan-Killiany atlas-based [23]) and 7 subcortical gray matter structures per hemisphere, lateral ventricle volumes, two whole-hemisphere measures and total intracranial volume were extracted, visually inspected and statistically evaluated for outliers (quality assurance is reported elsewhere [7]). Brain regions (features) used for classification included cortical thickness (CT), surface area (SA) and subcortical volumes of ROIs, two lateral ventricular and intra-cranial volumes (ICV), and two whole-hemisphere measures for SA and CT.

### Multivariate classification and validation

Participants with >10% missing entries were excluded (N=276), and median imputation was used for missing MRI data on the training set. Continuous features were centered around median zero and scaled according to their interquartile range. FS variables were combined with covariates age, sex, and site by concatenating individual feature vectors. All analyses were performed separately for pediatric and adult patients, and both groups combined. Common MVPA classifiers were applied: support vector machine (SVM) with linear and non-linear (radial-basis-function (RBF)) kernels, logistic regression (LR) with L1 and L2 regularization, Gaussian processes classification (GPC) with a linear kernel, and two decision-tree based ensemble methods, namely the random forest classifier (RFC) and the XGBoost algorithm [24–27]. A deep neural network was also implemented (fully connected; 3 layers with 60, 40 and 20 nodes respectively). SVM and LR classifiers were combined with and without automatic dimensionality reduction via principal component analysis (PCA), using the minimal number of components explaining 90% of the variance. Hyper-parameters for SVM (linear and non-linear), LR and XGBoost were optimized using nested cross-validation; RFC and GPC were tuned following recommendations. Details on handling missing data, model implementation and hyper-parametrization can be found in Supplementary **Methods**. The primary performance metric was area-under-the-receiver-operator-curve (AUC). Balanced accuracy, sensitivity and specificity are reported in supplement.

Multi-site classification of OCD patients versus HCs was assessed using different cross-validation (CV) approaches. First, we assessed multi-site classification using 10-fold CV to obtain maximally homogeneous train-test splits, with approximately the same number of subjects and the same proportion of samples coming from each site (internal validation). Next, we addressed leave-one-site-out (LOSO) CV, in which all but one site were used to train the models while the left out site was used to assess model performance (external validation). This may result in large between-sample heterogeneity of training and test sets, resulting in lower classification performance[28]. Because LOSO-CV has different fold sizes, we additionally performed 10-fold CV with LOSO-matched fold sizes, to evaluate whether differences in performance were due to differences in heterogeneity or fold size. Finally, we also performed single-site predictions using 10-fold CV to assess classification performance with reduced heterogeneity. Statistical significance of model performance was assessed directly through obtained AUC scores using the Mann-Whitney-U statistic for non-parametric testing (see supplement for details) [29]. Bonferroni-corrected level of significance was set at alpha=0.05 for the number of classifiers and comparisons.

### Clinical variables and sensitivity analysis

To explore the effects of clinical heterogeneity on classification performance, we selected subgroups with particular demographic and clinical characteristics: medication use, OCD severity, age of onset (AO) and duration of illness. Classifications performed were HC vs low (YBOCS<=24; mild-moderate[30]) and high severity (YBOCS>24; moderate-severe) OCD; HC vs early (<18yrs) and late AO (>=18yrs) OCD; HC vs short (<=7yrs) and long duration (>7yrs) OCD; and HC vs unmedicated and medicated OCD. For disease duration and severity, median splits were used to define groups; the 18 year threshold for AO was chosen in line with prior ENIGMA-OCD mega-analyses[6, 7]. Finally, as particular clinical variables can co-occur, we performed a post-hoc sensitivity analysis to investigate the effects of potential clinical covariance for results with AUC≥0.8 (see Supplementary **Methods**).

### Feature importance

To assess which brain regions and clinical variables contributed most to classification we used feature importance extracted from RFC combined with a permutation testing framework (see Supplementary **Methods**) [31].

## Results

### Multi-site classification

Three different CV approaches were used to assess the influence of sample heterogeneity. Results using various classification algorithms are summarized in **Figure 1**. Classification performance (AUC) using site-stratified CV (with training on combined samples and equal fold sizes) ranged between 0.57 (standard deviation (SD)=0.02;P_corr_=0.19) and 0.62 (SD=0.03;P_corr_<.001) across different classifiers. All models had statistically significant performance after multiple comparison corrections except for PCA+LR, PCA+SVM and NN classifiers. LOSO-CV led to lower classification performance; 0.51 (SD=0.11;P_corr_>.99) to 0.54 (SD=0.08;P_corr_>.99) AUC with relatively high variance across folds (SD=0.07-0.11) and no classifiers surviving multiple comparison corrections. AUC values obtained through site-stratified CV with different fold sizes were similar to site-stratified CV results with equal fold sizes, ranging between 0.57 (SD=0.08;P_corr_>.99) and 0.62 (SD=0.07;P_corr_=.55). However, variance across CV-iterations was higher and comparable to that from LOSO-CV (SD; site-stratified fixed: 0.02–0.04; site-stratified variable: 0.05-0.08; LOSO: 0.07-0.11). A complete overview of classification results is provided in supplementary **Table S2**. Multi-site classification, performed separately on pediatric and adult samples yielded similar results, ranging from 0.56 (SD=0.03;P_corr_>.99) to 0.62 (SD=0.06;P_corr_=.71) and 0.56 (SD=0.03;P_corr_=.69) to 0.61 (SD=0.02;P_corr_=.008) AUC, respectively (see supplementary **Tables S3-4**). As site-stratified CV with equal fold-sizes resulted in the best performances, we used this strategy for further evaluation of intra-site performance and the influence of clinical variables. Only RFC classification performance is reported here, as differences between classifiers were minimal and this model was also used to extract feature importance.

**Figure 1.**
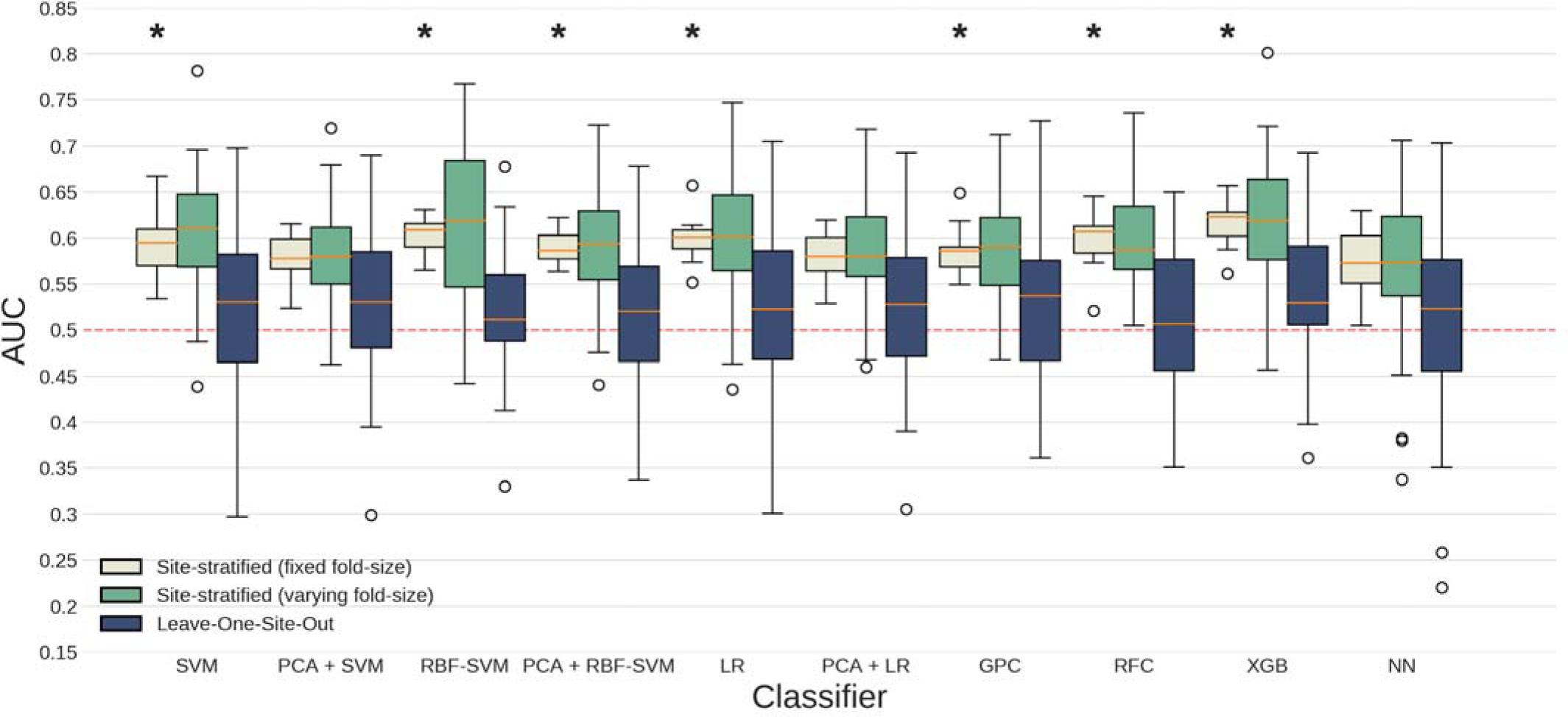
Performance for multi-site classification using different algorithms and cross-validation schemes. Boxplots summarize AUC scores obtained across CV-iterations; the dashed line represents chance-level performance and asterisks indicate scores significantly different from chance (Mann-Whitney-U statistic; p<0.05 Bonferonni corrected (10 classifiers x 3 CV types), see eSupplement for details). SVM=Support Vector Machine, PCA=Principal Component Analysis, RBF=Radial Basis Function, LR=Logistic Regression, GPC=Gaussian Processes Classification, RFC=Random Forest Classifier, XGB=XGBoost, NN=Neural Network.

### Single-site classification

Single-site classification performance with 10-fold CV varied greatly, with AUCs ranging between 0.17-0.91 across different sites and classifiers (see supplementary **Table S5**). **Figure 2** summarizes RFC performances for each individual site. We assessed the correlation between the number of participants and classification performance across sites, which showed a non-significant trend (r_S_=0.29, p=0.054).

**Figure 2.**
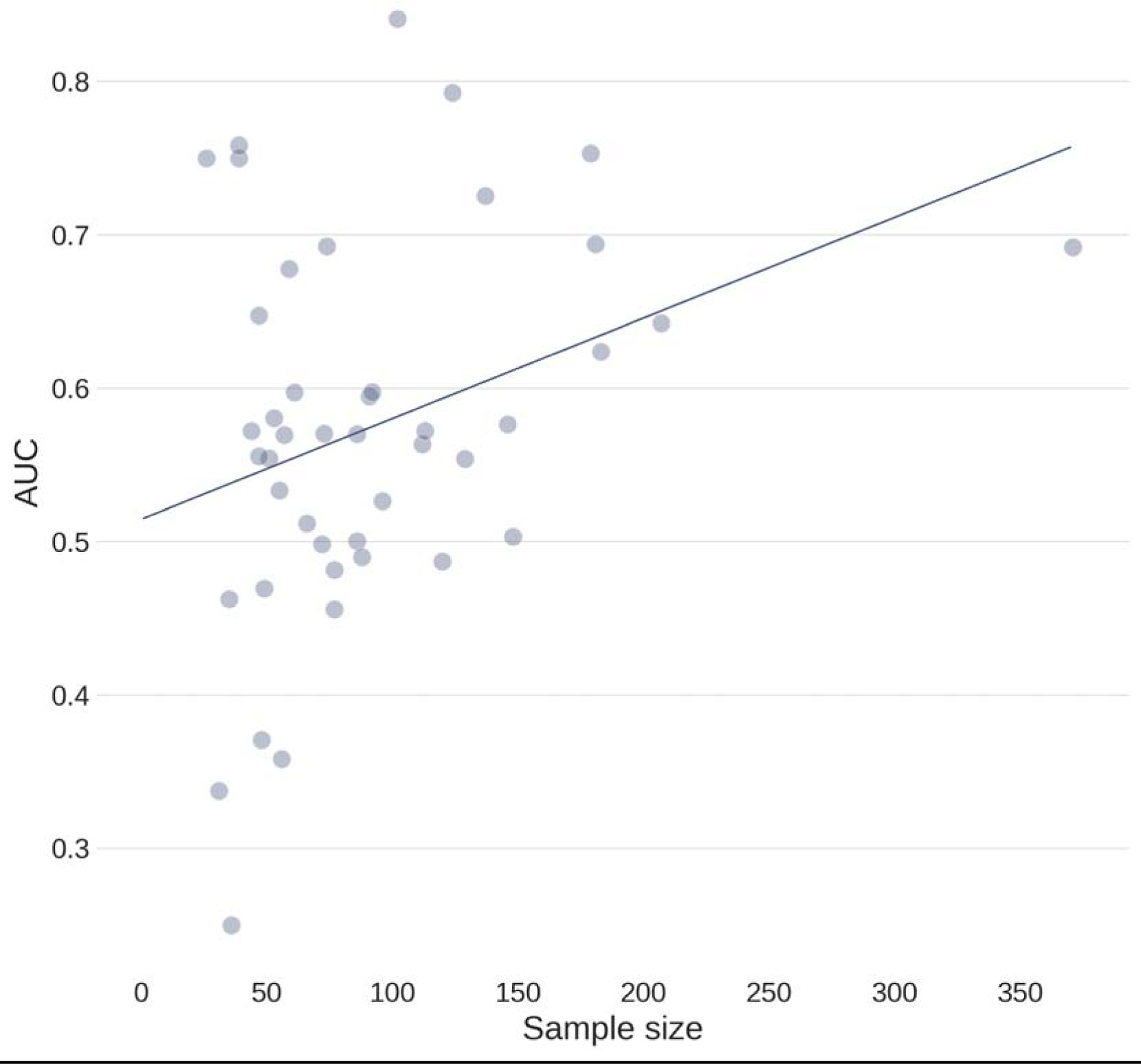
Scatterplot illustrating relationship between number of participants and classification performance across sites. Only RFC classifier performance averaged across CV-iterations are plotted (Spearman correlation; r_S_=0.29, p=0.054).

### Clinical variables and sensitivity analysis

To assess the influence of different clinical variables on classification performance, we repeated the analysis for specific subgroups split according to medication use, AO, disease duration, and severity. A complete overview is provided in supplementary **Tables S6(a-d)**, and we report results using RFC on combined data with age, sex and site as covariates below. Medicated OCD vs HC classification resulted in 0.73 AUC (SD=0.03;P_corr_<.001), unmedicated OCD vs HC in 0.61 (SD=0.02;P_corr_=.03), and medicated vs unmedicated OCD in 0.86 (SD=0.02;P_corr_<.001) (see **Figure 3**). Early AO OCD vs HC classification resulted in 0.68 AUC (SD=0.03;P_corr_<.001), late AO OCD vs HC in 0.73 (SD=0.02;P_corr_<.001), and early vs late AO in 0.81 (SD=0.03;P_corr_<.001). As no late AO patients were present in pediatric samples, classifications were re-run on adult samples only, resulting in 0.65 AUC (SD=0.04;P_corr_=.01) for early AO vs HC, 0.70 (SD=0.03;P_corr_<.001) for late AO vs HC, and 0.73 (SD=0.05;P_corr_<.001) for early vs late AO. Classification of short disease duration OCD vs HC resulted in 0.68 AUC (SD=0.04;P_corr_<.001), long disease duration vs HC in 0.71 (SD=0.02;P_corr_<.001), and short vs long duration in 0.78 (SD=0.04;P_corr_<.001). Finally, low severity OCD vs HC classification resulted in 0.60 AUC (SD=0.03;P_corr_=.15), high severity OCD vs HC in 0.61 (SD=0.03;P_corr_=.04), and low vs high severity OCD in 0.58 (SD=0.04;P_corr_>.99). Medication status correlated significantly with disease duration (r=-0.094; p<10^−05^; Bonferroni corrected). We therefore performed additional classifications after further stratification (e.g., HC vs medicated + short duration OCD; HC vs unmedicated + short duration OCD, etc). Classifications with or without stratification for disease duration were comparable (see supplementary **Tables S7(a-c)** for full overview).

**Figure 3.**
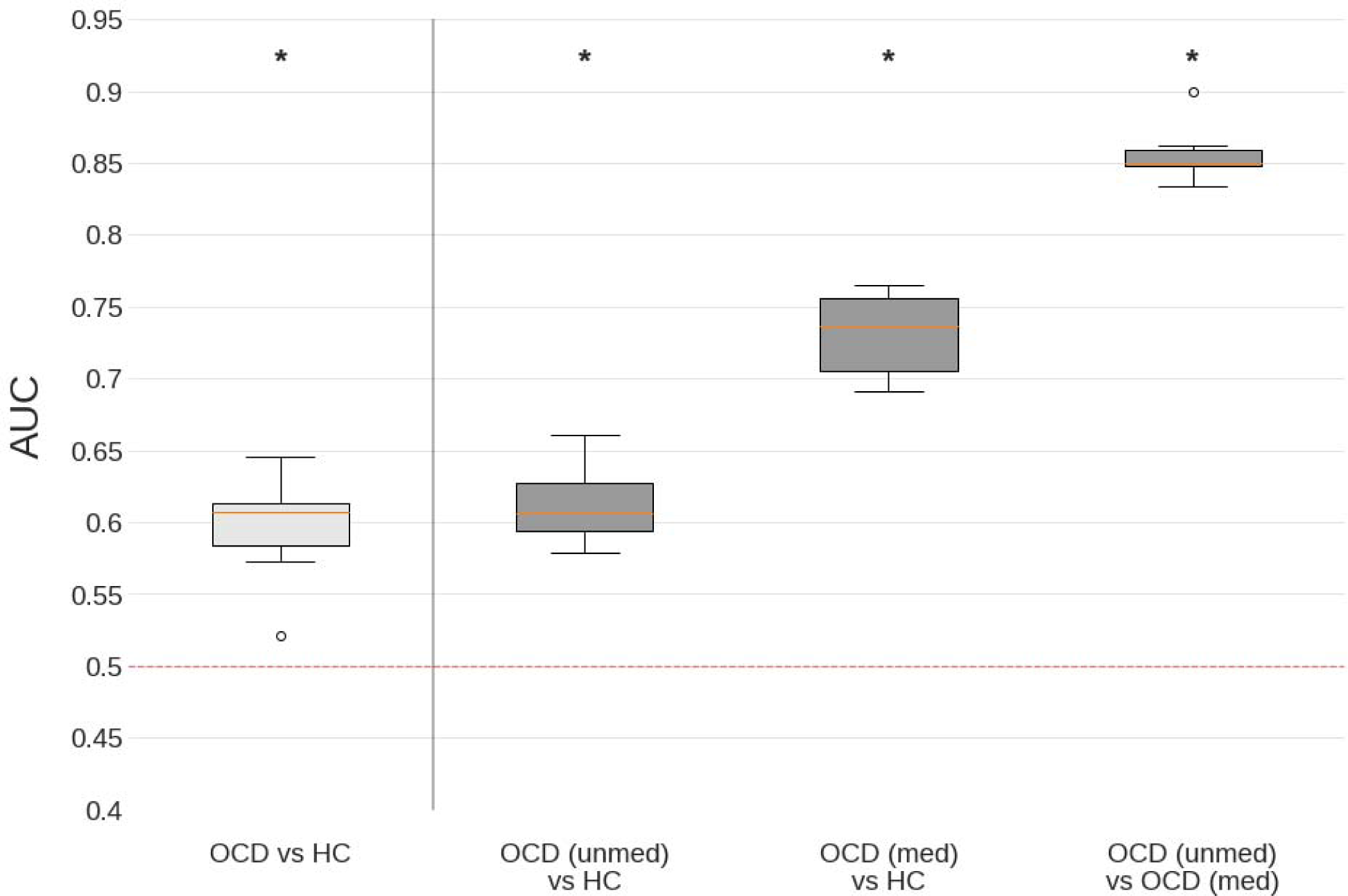
Performance for classification between subgroups of patients based on medication status. Only RFC classifier performance for combined data (both pediatric and adult samples) is shown here; Boxplots summarize AUC scores obtained across CV-iterations; the dashed line represents chance-level performance and asterisks indicate scores significantly different from chance (Mann-Whitney-U statistic; p<0.05 Bonferonni corrected (10 classifiers x 3 CV types), see eSupplement for details). unmed=unmedicated, med=medicated.

### Feature importance

We investigated which brain regions (features) contributed most to OCD vs HC classifications for site-stratified CV only, using the feature importance values from RFC and permutation testing. No features were selected consistently (survived false discovery rate (FDR) correction in >50% CV-iterations) for main analyses (OCD patients vs HC classification) in either pediatric, adult or combined samples. However, for HC vs medicated OCD and medicated vs unmedicated OCD classification in combined samples, 7 and 36 significant- and consistently selected features were found, respectively. Additionally, 40 features were found for both HC vs late AO and early vs late AO patients classifications in combined samples. A complete overview of these findings (including features importance for classifications stratified for medication and AO in adult samples) can be found in supplementary **Tables S8-11**.

## Discussion

We found that MVPA of parcellated structural neuroimaging data is unable to provide accurate distinction between OCD cases and controls. Classification of the complete sample using site-stratified CV ranged between 0.58 and 0.62 AUC, which is not sufficient for clinical application. Differences in performance between classifiers were minimal. Similar results were obtained for classifications performed separately on pediatric or adult samples. When validated on completely new data from other sites using LOSO-CV, model performance hardly exceeded chance-level. Our findings highlight the impact of validation schemes on classification performance and suggest poor discrimination between OCD patients and controls when combining data from multiple sites. In contrast, discrimination between subgroups of patients based on medication status enabled good individual subject classification.

Few diagnostic classifiers have been applied to OCD across multiple scanners and sites. Prior studies using structural MRI data to classify OCD using single-site samples yielded accuracies ranging from 0.72 up to 0.93 (reviewed in [12]). The wide range of performances observed in our individual site classification is in agreement with the published literature. Such a wide range may in part be explained by sample size, as larger samples tended to have higher AUC values [14, 28, 32]. However, this relationship does not necessarily hold true for large-scale multi-site studies, due to heterogeneity that arises from pooling samples with different scanning parameters, processing pipelines, inclusion criteria, demographic and clinical characteristics [12, 33]. All these factors can impact the data and obscure a pattern of abnormalities shared by all patients. Single-site studies that minimize heterogeneity may therefore yield higher classification performances, but limit the generalizability to new, unseen data and its use in clinical practice [14, 15]. LOSO-CV demonstrated that these structural MRI features do not provide a biomarker that enables generalization to new sites.

Classification within subgroups, split according to medication status, resulted in good performance even after accounting for correlated clinical variables (i.e., disease duration) through additional stratification. Evidence from rodent studies suggests that serotonin reuptake inhibitors (SRIs) mediate neuroplasticity in various cortical and subcortical structures through glio- and neuro-genesis [34–36]. However, little is known about how these findings might translate to humans and what the effects of long-term medication use are [37]. A few longitudinal studies suggest that SRI treatment normalizes brain volumes. One study reported significantly larger thalamic volumes in treatment-naïve pediatric patients compared to controls and these differences decreased following paroxetine treatment [38]. Another study reported smaller gray matter volume of CSTC-related regions in treatment-naïve patients that were no longer detectable following fluoxetine treatment [39]. Nonetheless, it remains unclear whether these structural changes are related to medication use or to symptom improvement.

Features that enabled multivariate classifications for medicated OCD vs HC included thickness of right medial orbitofrontal, right superior frontal, bilateral rostral middle frontal and right pars triangularis cortices in both adult and combined samples, and left palladic and right lateral ventricle volumes in adult samples only. Features for medicated vs unmedicated OCD in adult and combined samples included widespread cortical thickness in frontal, temporal, parietal and occipital regions. Although these multivariate features are important for the classifications as a whole, this appears consistent with previous univariate ENIGMA-OCD meta- and mega-analyses that also reported medication effects [6, 7]. Compared to HC, pediatric OCD patients had larger thalamic volumes and this finding was specific to unmedicated patients. This finding is in line with the normalizing effects of paroxetine on thalamic volume described previously. However, most findings have pointed towards more pronounced brain abnormalities in medicated patients than in unmedicated patients compared to controls. For example, medicated pediatric patients were found to have smaller cortical surface area (mainly in frontal regions) that was not detected in unmedicated patients. Medicated adult OCD patients showed thinner frontal, temporal and parietal cortices, and smaller hippocampal and larger pallidum volumes, whereas no differences were found for unmedicated adult patients. These results fit with the finding that the classification performance between medicated vs. unmedicated patients was better than that between cases versus controls, and may reflect minimization of heterogeneity in stratified patient groups compared to HC. Although these findings suggest that antidepressants affect brain structure that enables good single-subject classification, causal evidence for medication effects warrants prospective longitudinal studies. Furthermore, it remains unclear whether these findings are specific to OCD, or whether it is also present in other psychiatric disorders.

A number of limitations deserve emphasis. First, we used a sample pooled from existing data across the world, without harmonized protocols for scanning, inclusion criteria or demographic and clinical characteristics. These sources of heterogeneity may limit classification performance, but this also provides an opportunity for model development using independent data sets and the discovery of biomarkers that are reproducible across study sites. Second, limited information on medication use was available. We were therefore only able to distinguish patients on antidepressants with or without adjuvant antipsychotics versus those who had not received any medication. Medication history, medication dosage, and duration of use were unknown. Nonetheless, these coarsely defined medication groups enabled better case-control discrimination and good classification of medicated versus unmedicated cases. Third, there is a lack of information on comorbidity and OCD subtype in our dataset. Particular OCD subtypes may have different neural correlates, and this might limit the ability of MVPA models to find generalizable patterns in brain structure [12, 40]. Finally, it is possible that the brain features used for classification led to sub-optimal performance. OCD is thought to derive from abnormalities distributed at the network-level rather than focused on a single brain area, and FreeSurfer features might not be sufficiently sensitive to detect subtle alterations associated with OCD.

Taken together, this study provides a realistic estimate of the classification performance that can be achieved in a large, ecologically valid, multi-site sample of OCD participants using data on regional brain structure. Our findings show that parcellated structural MRI data does not enable a good overall distinction between patients with OCD and healthy controls. However, classifying subgroups of patients based on medication status enables good identification at the individual subject level. This underlines the need for longitudinal studies on the short- and long-term effects of medication on brain structure.

## Data Availability

Data are not publicly available.

## AUTHOR AND ARTICLE INFORMATION

Please address correspondence to W. B. Bruin. See the <u>online data supplement</u> for the complete list of ENIGMA OCD Working Group members.

